# Predicting Hospice Use Among American Indian/Alaska Native Persons with End-Stage Kidney Disease

**DOI:** 10.64898/2026.02.04.26345555

**Authors:** Brandon M. Varilek, Lauren E. Longacre, Farhad Shokoohi, Marcia Y. Shade, Prasanth Ravipati, Hossein Moradi Rekabdarkolaee

**Affiliations:** University of Nebraska Medical Center, College of Nursing – Omaha Division, 985330 Nebraska Medical Center, Omaha, NE 68198-5330, USA; University of Nebraska Medical Center, College of Nursing – College of Nursing, 985330 Nebraska Medical Center, Omaha, NE 68198-5330, USA; Department of Mathematical Sciences, University of Nevada Las Vegas, Las Vegas, NV 89154; University of Nebraska Medical Center, College of Medicine - Division of Nephrology Omaha, NE 68198, USA; Bowling Green State University, Department of Applied Statistics and Operations Research, Bowling Green, OH, USA

**Keywords:** predictive model, hospice, end-stage kidney disease, kidney transplant, American Indian/Alaska Native

## Abstract

**Background:** American Indian/Alaska Native individuals are disproportionately affected by end-stage kidney disease. Once diagnosed, treatment options include receiving a kidney transplant or starting dialysis. Disparities in treatment options are linked to social determinants of health, and in rural areas, dialysis is often the preferred treatment due to limited access to transplant. A recent nationwide analysis of survival differences showed a survival advantage after starting dialysis but also found that patients in these populations are less likely to receive hospice care before death. They face early diagnosis, leading to more years on dialysis and reduced quality of life. Using a predictive model can help identify factors that affect hospice use among patients. This study proposes a predictive model to estimate the likelihood of hospice use before death among patients who received a kidney transplant.

**Methods:** Using the 2022 USRDS Standard Analysis Files, adults who fit inclusion criteria were retrospectively identified. Area Deprivation Index (ADI) data, used for this analysis, includes 17 variables from US Census data, such as income, education, housing security, employment, and healthcare access, to generate the standardized ranking. This study employed regression tree, random forest, boosting, and support vector machine techniques to predict hospice use among participants. The covariates used in this study are race, IHS region, age, sex, mean ADI, comorbidities, and transplant status. Models are assessed based on their predictive performance using accuracy, sensitivity, and specificity.

**Results:** The random forest model outperforms others in accuracy, boosting offers the best sensitivity, and support vector machine and random forest excel in specificity. Overall, random forest and boosting are top models, with logistic regression as second-best. Logistic regression provides more interpretable results. The results, however, suggest a nonlinear relationship between covariates and the response that logistic regression might not capture. Based on both metrics, IHS region, age, and ADI are the most important features.

**Conclusions:** Including measures like the ADI in predictive models highlights geographic disparities that are often overlooked and can guide interventions toward communities that have been historically underserved. Researchers and healthcare systems should improve access to hospice and palliative care for those who face these disparities.

## BACKGROUND

American Indian/Alaska Native (AI/AN) persons are disproportionately affected by end-stage kidney disease (ESKD) with a prevalence rate 2.4 times that of non-Hispanic Whites.^1-3^ Furthermore, AI/ANs are two to three times as likely to be diagnosed with diabetes than non-Hispanic Whites and nearly two-thirds of all ESKD is caused by diabetes (ESKD-D) in AI/AN communities.^4-6^ Once diagnosed with ESKD, options for continued survival include receiving a kidney transplant or initiating dialysis.^7^ Dialysis and transplant are both financially taxing. Annual dialysis-related expenses average $86,400 per person.^8^ Furthermore, kidney transplant and the life-long immunosuppressant drug therapies are also expensive (average surgery cost is $442,500;^9^ average standard drug therapy cost is $25,000 per year^10^).

For AIs who reside in rural/frontier areas (like South Dakota, United States), dialysis is often the treatment of choice, as transplant access is severely limited. The inequity in treatment modalities is tied to social determinants of health (SDOH), such as chronic underfunding of healthcare services,^11-13^ racial discrimination,^14-16^ geographic placement onto reservations,^17,18^ and extensive poverty.^19,20^ Though dialysis is life-prolonging, quality of life (QOL) can be significantly impacted by the repeated treatment regimen.^21^ Integrating palliative care into the dialysis setting has the potential to address negative QOL in AI/ANs with ESKD. However, access to palliative care use in dialysis settings is rare^22^ and palliative care access for AI/ANs in general remains elusive.^23^ Additionally, it is difficult to assess the rate of palliative care referrals and use for those with ESKD due to inconsistent use of diagnostic codes. However, data regarding the use of hospice prior to death is reported directly to the United States Renal Data System (USRDS) at the time of death.

A recent nationwide analysis of survival disparities between AI/ANs and NHW revealed that AI/ANs have a survival advantage over NHW after starting dialysis^24^ and AI/ANs are significantly less likely to receive hospice prior to death.^25^ However, AI/ANs are diagnosed much earlier than NHWs, leading to additional years on dialysis and greater potential for poor QOL.^26^ SDOH were shown to play a significant role in the survival models especially as SDOH metrics worsened. However, there appeared to be a non-linear behavior between the covariates and the response in the modeling that a linear logistic regression cannot capture, especially when the Area Deprivation Index® (ADI®) is used in correlation with datasets with differing geospatial resolution. What remains unknown is how SDOH impact the likelihood of receiving hospice prior to death while receiving dialysis, valuable data that could enable researchers to developed targeted interventions for those who are currently least likely to receive hospice. The use of a predictive modeling approach may help better describe the non-linear nature of SDOH and hospice use in AI/AN populations with ESKD.

Predictive models aid researchers and clinicians in making more informed decisions by providing insights into future trends, outcomes, or events based on historical data.^27^ Furthermore, predictive modeling allows organizations to optimize resource allocation by identifying areas where resources can be deployed more efficiently.^28^ For example, it is well known that palliative and hospice care is challenging for AI/ANs to obtain due to SDOH.^11-20^ Using a predictive model can provide healthcare clinicians and researchers with a “profile” of common characteristics that would lead someone with ESKD to use hospice prior to death and a profile of characteristics of someone who would not receive hospice. These profiles can point researchers towards intervention development for populations who are most at risk of not receiving hospice prior to death. Additionally, predictive modeling strategies help identify the most important variables that make up the model and are driving the model outcome.

For this study, our aim is to develop a predictive model with hospice use as the response variable using USRDS and SDOH data to accurately predict the likelihood receiving hospice prior to death among AI/ANs and NHWs who have received a prior kidney transplant.

## METHODS

This manuscript reports the findings of a secondary data analysis using USRDS data. Data used with permission of the USRDS under a completed data use agreement. The study was approved by the South Dakota State University Institutional Review Board.

### Cohort Derivation

Using the 2022 USRDS Standard Analysis Files,^29^ all adults aged ≥ 18 years from the main PATIENTS file who initiated chronic hemodialysis or peritoneal dialysis after January 1, 2014, were retrospectively identified for inclusion for this predictive model development. Patients who did not identify as AI/AN or NHW were excluded from the analysis. Patients were further limited to only include the “diabetes” primary disease diagnosis group, which included all relevant primary diabetes-related ESKD ICD-9 and ICD-10 codes found on the CMS-2728 forms.^30^ Patients with a history of kidney transplant were included in the analysis. The DEATH file was used to identify those who had died on January 1, 2000, or after. Data was merged with the main PATIENT data using the USRDS ID. Hospice was the key indicator for this analysis and is in the DEATH file and reports hospice use as yes, no, or unknown. Only patients with yes or no were included in the predictive model. A total of 317,577 patient records were included in the model development.

### Social Determinants of Health Data and Data Fusion

The Neighborhood Atlas^®^ publishes the ADI^®^ that ranks neighborhoods by socioeconomic disadvantage.^31^ ADI^®^ data providing national comparisons were used for this analysis. The ADI^®^ considers 17 material and socioeconomic variables gathered by US Census data^32^ to provide the standardized ADI^®^ ranking and includes income, education, housing security, employment status, and healthcare access.^33,34^ Neighborhood Atlas^®^ reports ADI^®^ by census block groups from 1 to 100; a rating of 1 represents the lowest level of disadvantage and a score of 100 represents the highest level of disadvantage.^35^ The ADI^®^ gives an estimate of the impact of SDOH in a particular neighborhood.

The national level ADI^®^ data is provided at 9-digit zip code level (5-digit zip code first followed by 4-digits to identify specific location information [12345-6789]^36^). However, USRDS data only contains 5-digit zip codes which limits the usefulness of the 9-digit zip code. As an example, a single 5-digit zip code can include several hundred 9-digit census block units with ADI^®^ ratings that span anywhere from 1 to 100, making it impossible to correlate the 5-digit zip code with the proper ADI^®^ rating. Therefore, we upscaled all 9-digit zip codes by taking the average of all 9-digit zip codes that had the same 5-digit prefix. This process was carried out for all ADI^®^ data. Then, the average ADI^®^ for each 5-digit zip code level was attached to the original USRDS data using the zip code variable.

### Predictive Model Development

Logistic regression is the standard statistical model used for analyzing and predicting binary response using a set of predictors. This approach is easy to interpret in terms of using the odds ratio. Despite its popularity, logistic regression can only model relationships that are linearly separable in the log-odds space. Therefore, if the true relationship is highly nonlinear, the predictive performance of the model drops unless one engineer features or use polynomial terms. To address this, many machine learning (ML) models have been developed to improve prediction accuracy. Although these models are capable to capture the nonlinear patterns in the data, their outputs are often not easy to interpret. Furthermore, they are prone to overfitting without regularization and often computationally expensive than parametric models like logistic regression. This study employed regression tree (RT), random forest (RF), boosting, and support vector machine (SVM) to predict hospice use among AI persons with a previous kidney transplant. The RF is a machine learning (ML) algorithm for classification based on the recursive partitioning principle. For RF, specific information about the relationships between the response and predictor variables is not required.^37-39^ It creates a forest with several decision trees. With the RF approach, the accuracy and robustness of the model is directly correlated with the number of trees in the forest.^37^ We experimented with various numbers of trees and discovered that 50 trees produced the most precise predictions. The Support Vector Machine (SVM) creates a line or a hyperplane that separates the data into different classes. The line or hyperplanes are considered as the decision boundary, and they are used to predict continuous outputs. The covariates used in this study are race, IHS region, age, sex, mean ADI, and comorbidities. Figure 1 shows a graphical representation of the general form of each model employed in this study.

**Figure 1.**
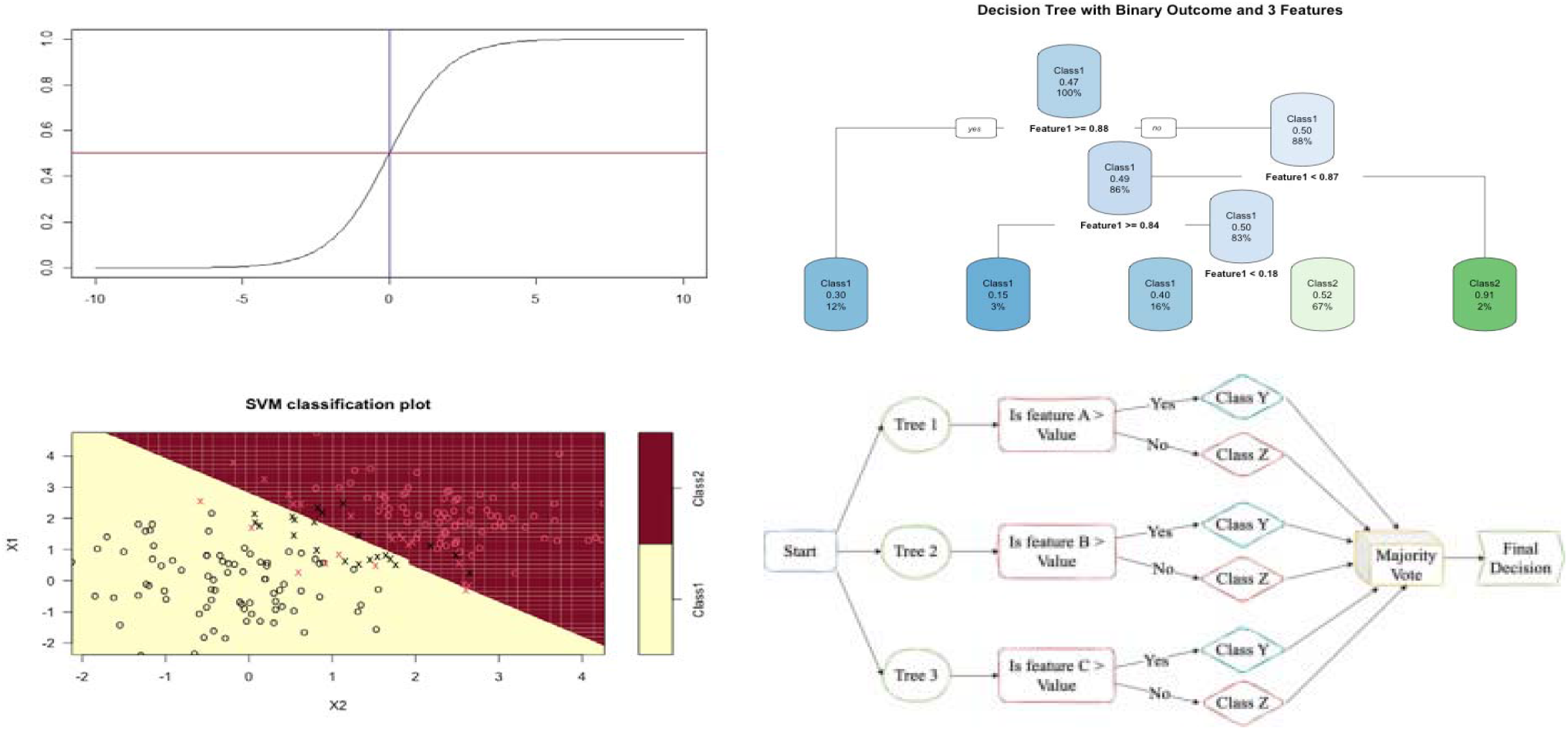
General form of logistic regression (Top Left), Decision Tree with 3 features (Top Right), Support Vector Machine with linear kernel (Bottom Left) and random forest resulting from combining 3 decision Tree (Bottom Right).

### Statistical Analysis

All the data analysis in this study were conducted using R 4.2.2 at the 95% confidence interval (α = 0.05). Models are assessed based on their predictive performance using accuracy, sensitivity, and specificity. Accuracy is determined by the ratio of correct predictions to the total number of predictions made. Sensitivity, or the true positive rate, represents the likelihood of a positive test result given that the individual is truly positive. On the other hand, specificity, or the true negative rate, indicates the probability of a negative test result given that the individual is truly negative. Additionally, prevalence serves as a measure of disease, reflecting the likelihood of an individual having the condition. It is calculated by multiplying the incidence rate by the average duration of the disease. Higher prevalence may suggest prolonged survival without a cure or an increase in new cases, while lower prevalence may indicate more deaths than cures or fewer new cases. Prevalence rises with the identification of new cases and decreases when a patient is cured or passes away. It’s important to note that a disease can exhibit high prevalence and low incidence, or vice versa.

## RESULTS

Figure 2 shows the results of each model for the cumulative sample for each year and after, and Figure 3 shows the results for each of the years alone. Each figure displays the results for each model. The higher accuracy, sensitivity, and specificity denote better performance. Based on these plots, the random forest model outperforms other methods in terms of accuracy, Boosting provides the best sensitivity, and Support vector machine and random forest provides are best in terms of specificity. Overall, Random Forest and Boosting seem to be best models and logistic regression to be the second-best model and provides results that are very close to the machine learning models. Furthermore, logistic regression provides an interpretable result compared to the machine learning models. The results suggest that there might be a nonlinear relationship between the covariates and response variable that logistic regression was not able to capture.

**Figure 2.**
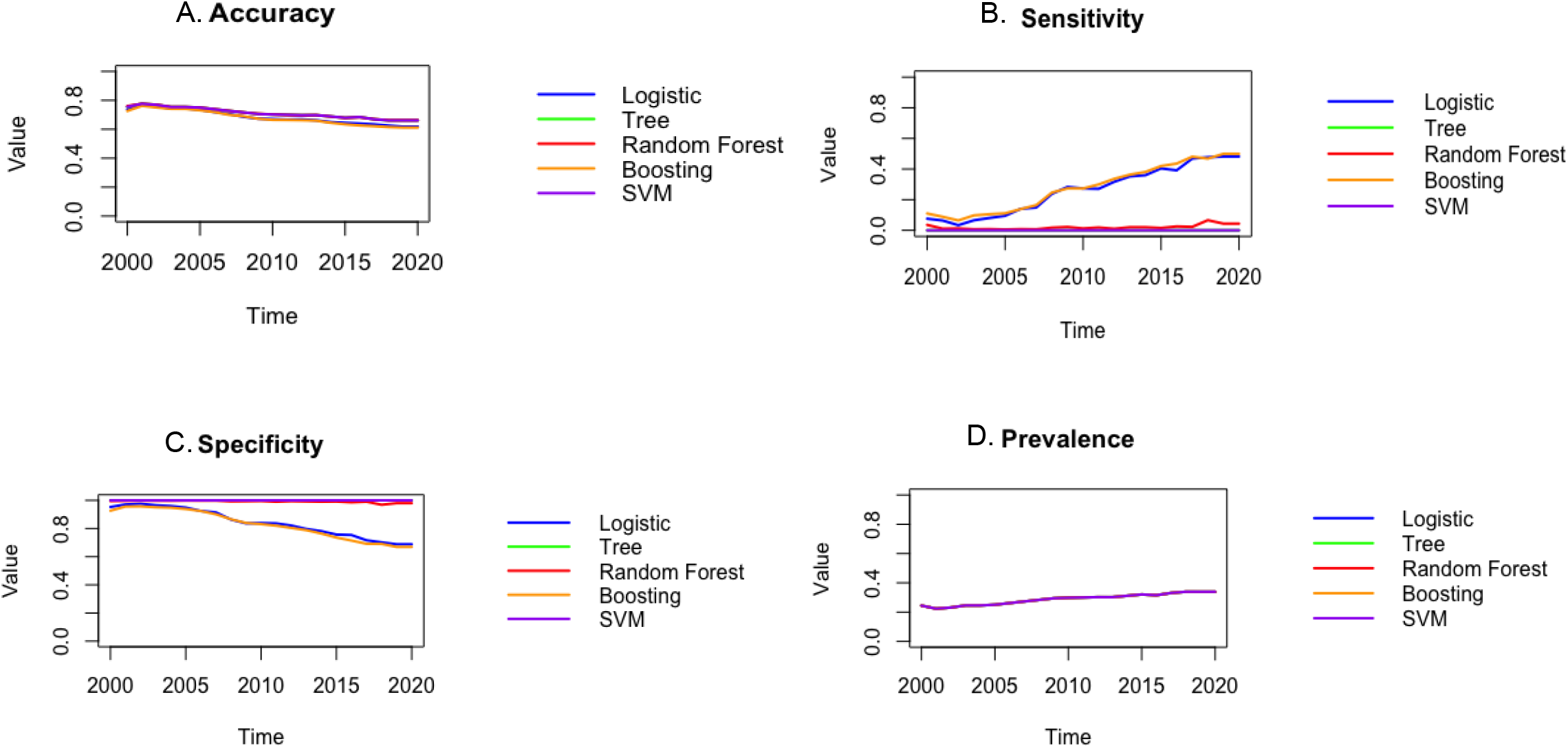
The accuracy (A), sensitivity (B), specificity (C), and prevalence (D) plots for the cumulative data for each year and after that. The higher accuracy, sensitivity, and specificity denote better performance. Based on these plots it can be observed that the random forest model outperforms other methods in terms of accuracy, sensitivity, and specificity. Furthermore, boosting seems to be the second-best model.

**Figure 3.**
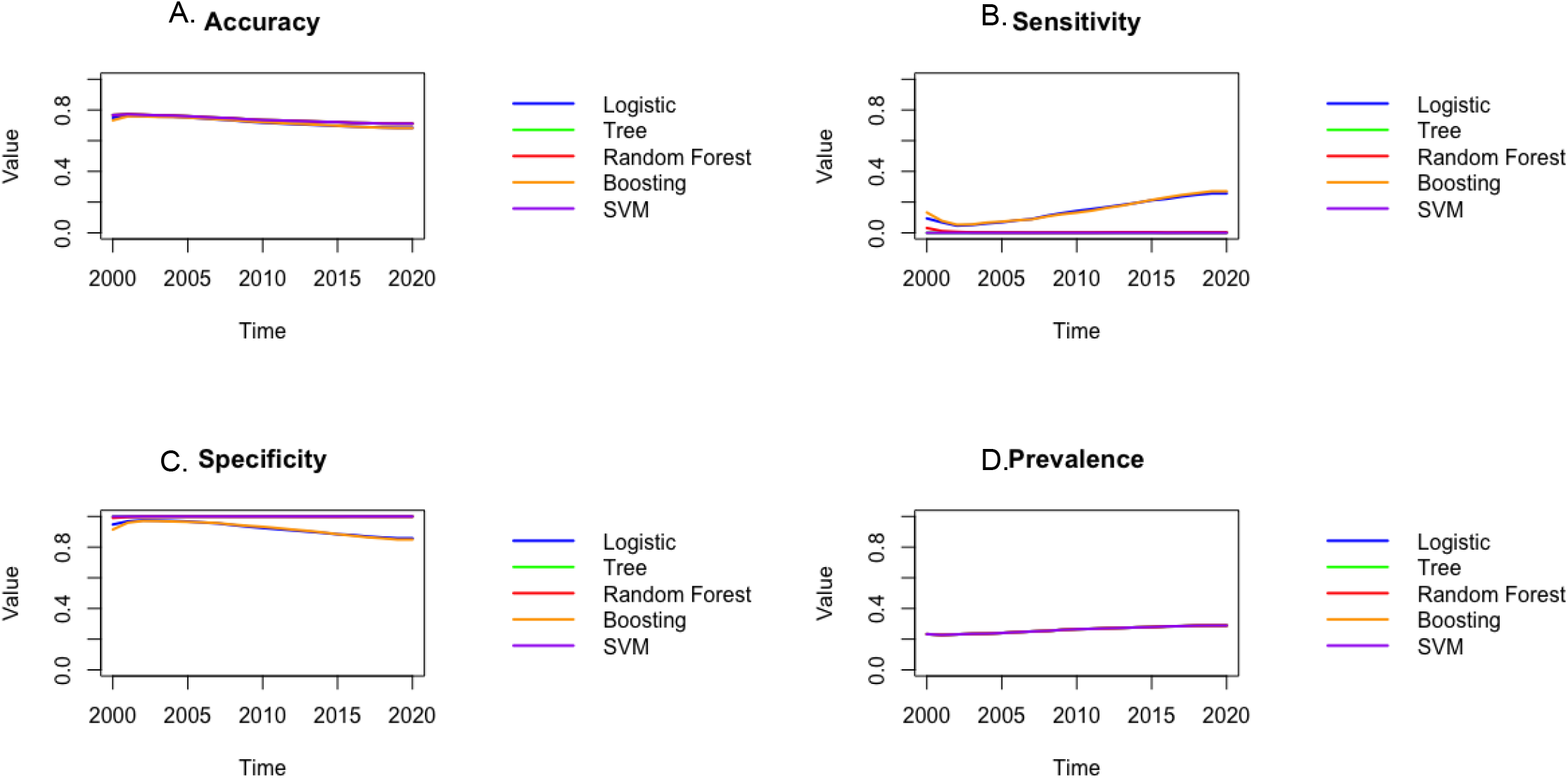
The accuracy (A), sensitivity (B), specificity (C), and prevalence (D) plots. The higher accuracy, sensitivity, and specificity denote better performance. Based on these plots it can be observed that the random forest model outperforms other methods in terms of accuracy, sensitivity, and specificity. Furthermore, boosting seems to be the second-best model.

Figure 4 shows the post hoc analysis of each of the 3 top models. The two commonly used metrics for feature importance in Random Forests are Mean Decrease Accuracy (MDA) and Node Purity also called Mean Decrease Gini (MDG). The MDA, also known as permutation importance, assesses the impact of each feature on model performance by measuring the decrease in prediction accuracy when the feature values are randomly shuffled. The Node Purity measures the total increase in Gini purity when a feature is used to split data within the trees. The Node Purity is efficient to compute and offers a fast way to rank features, but it tends to overemphasize features with many categories or higher cardinality and does not directly reflect their impact on model performance.^37,40,41^ Based on both metrics, IHS region, age, and ADI are the most important features. Similar results have been found using the relevant importance for the boosting method. Although history of a kidney transplant significantly reduces the likelihood of receiving hospice prior to death, it was relatively insignificant in the variable importance plots. For the logistic regression, the results shows that the odds ratio for the race, sex, IHS Region 12, and age of diagnosis are significantly away from 1. Among these, only the odds ratio of race is greater than 1, indicating an increased odds of race predicting hospice use. For the other variables, the odds ratios are less than 1 which indicates a decrease in the odds of hospice use.

**Figure 4.**
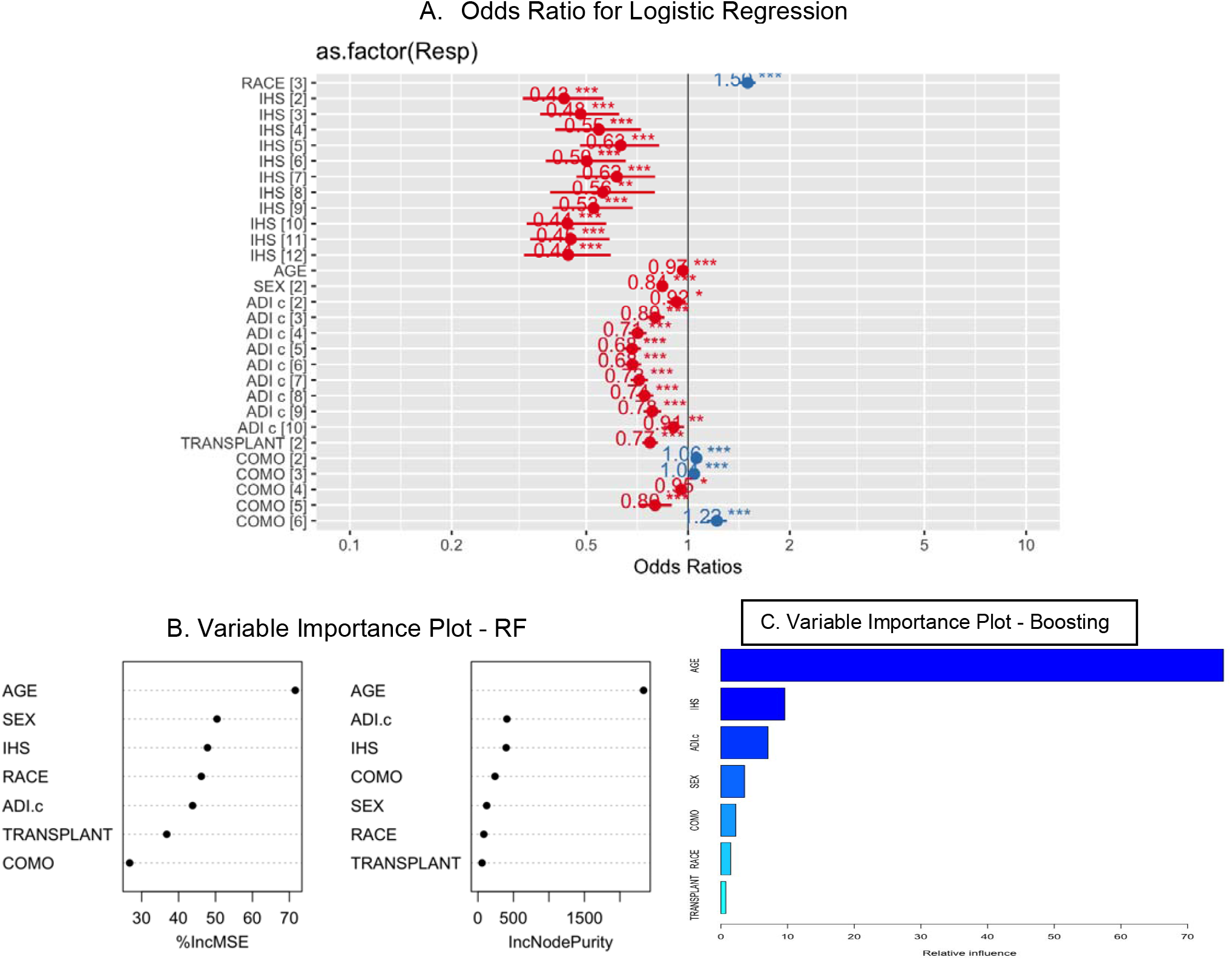
The plot of the odds ratio for logistic regression (A), important variable in Random Forest (B), and relative importance plot for boosting (C). For Logistic regression, race, IHS 12, diagnostic age, and sex are significant. For the Random Forest model, IHS, diagnostic age, and ADI seem to be the most important variable. For the Boosting model, ADI, diagnostic age, and IHS are the most important variables.

## DISCUSSION

This study reports our preliminary predictive model of USRDS and SDOH data to predict the likelihood of AI/ANs and NHWs with ESKD-D to receive hospice prior to death. In the RF model, the most important variables were IHS Region, age at diagnosis of ESKD-D, and ADI® rating. These variables are also the most important variables for the Boosting model, but the importance of the variables appears in a different order. Despite the importance of these variables in the RF and Boosting models, these three variables are not significant in the logistic regression. This could indicate that there is a complex relationship between the variables (geographic location, age at diagnosis, ADI® rating, and hospice use prior to death) that is not linear.

This predictive model for hospice use is a first step at developing an improved predictive model that can account for longitudinal patient data, a predictive model that has not been previously developed in persons with ESKD-D. Since USRDS data only provides patient baseline and death data for this sample, the application of the results may be limited (static prediction modeling^42^). This means that changes in patient condition over time are not accounted for, including changes to SDOH. Further, disease progression, care preferences, and service use over time cannot be assessed. Given the time-varying nature of this problem, it is essential that we account for longitudinal data to provide a dynamic prediction model. Dynamic modeling has been used in persons with chronic kidney disease,^42^ yet SDOH variables were not integrated into the model, leaving gaps in the prediction outcomes that need to be addressed.

The Center for Medicare and Medicaid Services (CMS) developed their Kidney Care Choices (KCC) model^43^ that has improved the quality of kidney care, increased home dialysis use, and has provided pathways that allows patients to receive dialysis or transplant services concurrently with hospice services.^44^ Developing a predictive model that provides healthcare clinicians with a “profile” of characteristics that may reduce an individuals’ likelihood of receiving hospice care would not only bolster the outcomes of the KCC, but provide an opportunity for more people with ESKD-D to access hospice services. SDOH greatly limit AI/ANs access to hospice services, so a predictive model highlighting those most at risk for not receiving hospice can enable interdisciplinary clinicians to assist people in overcoming health-related social needs that make hospice access challenging. Based on these results, ADI®, race, and age of ESKD diagnosis are the most important factors in predicting the likelihood of receiving hospice. Therefore, researchers and healthcare systems must strive to improve hospice and palliative care access to AI/ANs and NHWs who live in areas where negative effects of SDOH are prevalent.

Beyond improving the existing model with longitudinal data, there is value in examining other predictive frameworks within palliative and hospice care—particularly those developed to anticipate service use. Kawashima et al.^45^ demonstrated the feasibility of using machine learning to predict specialist palliative care needs among advanced cancer patients, leveraging a small set of longitudinal features. Similarly, Murphree et al.^46^ implemented a real-time model integrated within clinical workflows in a hospital setting to identify inpatients most likely to benefit from palliative care consultation. Notably, both models prioritized the prediction of service use, as opposed to relying on outcomes such as mortality, relevant to our own approach.

### Limitations

Our study is not without limitations. The development of this predictive model is based persons with a diagnosis of ESKD-D as found within the CMS Medical Evidence Report (CMS Form 2728) at the time of dialysis initiation.^30^ Therefore, predictions of hospice use in persons with ESKD-D cannot be extrapolated to broader diagnoses of ESKD without inputting addition data into the model. Additionally, the USRDS data used in this work is only captured at dialysis initiation and at death. It is possible that there are other variables present in electronic medical record systems that could provide more accurate predictions of hospice use, but this exceeded the scope of this project. Future work should incorporate a wider array of variables to improve the accuracy and sensitivity of the predictive model.

## CONCLUSION

When predictive models include measures such as the Area Deprivation Index (ADI), they can help expose geographic patterns of inequity that might otherwise go unnoticed. This kind of insight is particularly important for targeting interventions in communities that have been historically underserved by hospice and palliative care. Therefore, researchers and healthcare systems must strive to improve access to hospice and palliative care for AI/ANs and NHWs, who are disproportionately affected by these inequities.

## Supporting information

USRDS Manuscript Approval

## Data Availability

Restrictions apply. Data used for this project were obtained from USRDS via a data use agreement and are not able to be shared in raw formats. Data for ADI values are available through the Neighborhood Atlas website

https://www.neighborhoodatlas.medicine.wisc.edu/

https://www.niddk.nih.gov/about-niddk/strategic-plans-reports/usrds/for-researchers/standard-analysis-files

## LIST OF ABBREVIATIONS

ADI®: Area Deprivation Index®
AI/AN: American Indian/Alaska Native
ESKD: End-stage kidney disease
ESKD-D: End-stage kidney disease related to diabetes
ML: Machine learning
NHW: non-Hispanic White
QOL: Quality of life
RF: Random forest
RT: Regression tree
SD: South Dakota
SDOH: Social determinants of health
SVM: Support vector machine
USRDS: United States Renal Data System

## ACKNOWLEDGMENTS

The data reported here have been supplied by the United States Renal Data System (USRDS). The interpretation and reporting of these data are the responsibility of the author(s) and in no way should be seen as an official policy or interpretation of the U.S. government.

## AVAILABILITY OF DATA AND MATERIALS

Restrictions apply. Data used for this project were obtained from USRDS via a data use agreement and are not able to be shared in raw formats. Data for ADI^®^ values are available through the Neighborhood Atlas^®^ website: https://www.neighborhoodatlas.medicine.wisc.edu/

---

**Table 1.** Cohort Characteristics.

|  | Previous Kidney Transplant |  |  | No Prior Kidney Transplant |  |  |
| --- | --- | --- | --- | --- | --- | --- |
|  | Total (N) | AI/AN (n [%]) | NHW (n [%]) | Total (N) | AI/AN (n [%]) | NHW (n [%]) |
| **Nationwide Sample** | ~8,914<sup>†</sup> | ~363 (4.1)<sup>†</sup> | 8,551 (95.9) | 308,663 | 9,643 (3.1) | 299,020 (96.9) |
| **Gender (%)** |  |  |  |  |  |  |
| Female | 3,149 (35.3) | 155 (42.7) | 2,994 (35.0) | 132,084 (42.9) | 4,962 (51.5) | 127,122 (42.5) |
| **Age (at ESKD Diagnosis-in years)** |  |  |  |  |  |  |
| 18-44 | 2,243 (25.2) | 71 (19.5) | 2,172 (25.4) | 15,621 (5.1) | 1,175 (12.2) | 14,545 (4.9) |
| 45-54 | 2,414 (27.1) | 118 (32.4) | 2,296 (26.9) | 35,979 (11.7) | 2,072 (21.5) | 33,907 (11.3) |
| 55-64 | 2,953 (33.1) | 131 (35.9) | 2,822 (33.0) | 77,948 (25.3) | 2,980 (30.9) | 74,968 (25.1) |
| 65-74 | 1,221 (13.7) | 43 (11.8) | 1,178 (13.8) | 97,493 (31.6) | 2,384 (24.7) | 95,109 (31.8) |
| 75+ | ~83<sup>†</sup> (0.9) | <11 | 83 (1.0) | 81,523 (26.4) | 1,032 (10.7) | 80,491 (26.9) |
| **Hospice Use** |  |  |  |  |  |  |
| Yes | 2,202 (24.7) | 80 (22.0) | 2,122 (24.8) | 89,560 (29.0) | 1,922 (20.0) | 87,638 (29.3) |
| No | 6,712 (75.3) | 283 (78.0) | 6,429 (75.2) | 219,103 (71.0) | 7,721 (80.0) | 211,382 (70.7) |
| **Comorbidities (N/n with comorbidity present at dialysis start date)\*** |  |  |  |  |  |  |
| AHD | 1,522 | 63 | 1,459 | 74,954 | 1,626 | 73,328 |
| Cancer | 562 | 25 | 537 | 23,871 | 489 | 23,382 |
| CHF | 2,404 | 119 | 2,285 | 138,281 | 3,441 | 134,840 |
| CVA/TIA** | 901 | 38 | 863 | 39,575 | 1,041 | 38,534 |
| Tobacco User | 722 | 24 | 698 | 25,102 | 848 | 24,254 |
| **Total Number of Additional Comorbidities (n (% of nationwide n))\*** |  |  |  |  |  |  |
| 0 |  | 198 (54.5) | 4,907 (57.4) |  | 4,856 (50.4) | 111,601 (37.3) |
| 1 |  | 115 (31.7) | 2,438 (28.5) |  | 3,040 (31.5) | 111,197 (37.2) |
| 2 |  | 29 (7.9) | 790 (9.2) |  | 1,269 (13.2) | 56,051 (18.7) |
| 3 |  | <11 | 119 (1.4) |  | 243 (2.5) | 13,797 (4.6) |
| 4 |  | <11 | 13 (0.2) |  | 32 (0.3) | 1,487 (0.5) |
| 5 |  | 15 (4.1) | 283 (3.3) |  | 202 (2.1) | 4,740 (1.6) |

\*Data available from CMS-2728 Form Box 16 (co-morbid conditions).<sup>30</sup> AI/AN: American Indian/Alaska Native; AHD: Atherosclerotic Heart Disease; CHF: Congestive Heart Failure; \*\*CVA/TIA includes history of cerebrovascular accident (CVA) and transient ischemic attack (TIA); ESKD: End-stage kidney disease; NHW: Non-Hispanic White.
<sup>†</sup>Total number of additional comorbidities include any combination of the 5 comorbid variables included in the analysis; <sup>†</sup>Approximate values given per USRDS guidelines for <11 identified eligible patients in a cell.

