## Supplementary material for "Predicting Hospice Use Among American Indian/Alaska Native Persons with End-Stage Kidney Disease": USRDS Manuscript Approval

**DEPARTMENT OF HEALTH & HUMAN SERVICES**

Public Health Service
National Institute of Health

National Institute of Diabetes and
Digestive and Kidney Diseases
Bethesda, Maryland 20892

February 2, 2026

Kimberly Nieman
United States Renal Data System
701 Park Ave S4.100
Minneapolis, MN 55415

Re: USRDS Manuscript Approval # MS2026-03

Dear Kimberly:

I have reviewed the manuscript entitled, "Predicting Hospice Use Among American Indian/Alaska Native Persons with End-Stage Kidney Disease", submitted by Dr. Brandon Varilek on behalf of Dr. Hossein Moradi Rekabdarkolaee of South Dakota State University. The manuscript is based on DUA 2022-55b.

This manuscript fulfills USRDS privacy requirements and is manuscript MS2026-03.

Please wish the authors good luck with the submission. I would appreciate being notified if or when the manuscript is accepted for publication.

Sincerely,

Tracy L. Rankin, PhD, MPH
Program Official
National Institute of Diabetes and Digestive and Kidney Diseases
National Institutes of Health
Bethesda, Maryland 20892
